# Assessing potential harms from screening overdiagnosis and false positives with multicancer early detection tests

**DOI:** 10.64898/2026.04.09.26348927

**Authors:** Talía Malagón, W. Alton Russell, Julia V. Burnier, Kyle Dickinson, Darren Brenner

## Abstract

**Background:** Multicancer early detection tests could be used for cancer screening, but may lead to harms, including false positive results and overdiagnosis of indolent tumours that would not have become clinically evident during that person’s lifetime. We assessed the potential for these screening harms in the context of future population-based screening with a multicancer early detection test.

**Methods:** We used a microsimulation model to assess potential population-level impacts of screening at ages 50-75 years with a multicancer early detection test in Canada. We assumed high test specificity (97-99.1%) and test sensitivity increasing with cancer stage. The model includes latent indolent cancers that would not be diagnosed within that person’s lifetime but can be overdiagnosed through screen-detection. We calculated the yearly and cumulative lifetime probabilities of screening overdiagnosis and false positive test results, assuming a range of preclinical screen-detectable periods (2-5 years).

**Results:** An estimated 2.1-6.0% of all yearly screen-detected cancers with a multicancer screening test were predicted to be overdiagnoses across scenarios. The proportion of overdiagnosis varied by site, and strongly increased with age, going from 1% at age 50 to over 10% of screen-detected cancers by age 75. The test positive predictive value ranged from 15.9%-77.6%, meaning that there could be 0.3-5.3 false positives with no underlying cancer for every true cancer case detected by the test.

**Conclusion:** Population-level multicancer screening with a multicancer early detection test would likely not lead to substantial screen-related overdiagnosis. Healthcare systems should consider how screening false positives may increase their diagnostic service caseload.

## Introduction

Multicancer early detection tests are emerging technologies that seek to identify cancer at multiple sites using a single blood test, and which could be used for population-based screening.^1,2^ These contrast with prior tests used for population-based cancer screening, which only test for cancer at a single site. Most multicancer tests currently available or in development are based on the examination of mutation and methylation patterns in cell-free DNA and circulating tumour DNA (ctDNA), but may also assess other biomarkers such as proteins.^1^ While these tests are not currently recommended for cancer screening, clinical studies have provided proof of concept that they are able to identify cancers in asymptomatic populations.^3,4^ Randomized controlled trials of these tests for screening are either currently underway or in the planning stages.^5,6^ If eventual trials show these tests are effective at reducing cancer late-stage diagnosis and mortality, then policy makers will need to seriously consider whether these should be recommended for multicancer screening.

Decisions on whether to recommend a test for population-based cancer screening are based on an assessment of whether a screening test’s benefits outweighs its harms. While earlier cancer detection may reduce cancer morbidity and mortality, the potential harms of screening include false positives, unnecessary confirmatory tests, and screening overdiagnosis. Overdiagnosis is defined as an indolent cancer detected through screening that would not otherwise have been diagnosed within that person’s lifetime, as the person would have died from other causes prior to the cancer becoming clinically apparent.^7^ Overdiagnosed cancers represent an important harm of screening, as these patients undergo unnecessary and potentially harmful interventions that do not extend their lifespan, and contribute to healthcare system overcrowding, delaying care for other patients. The potential for overdiagnosis with multicancer early detection tests has been understudied, with the risk often dismissed as negligible due to the high prognostic vale of ctDNA.^8^ However, experience with previous cancer screening programs has shown that the introduction of a new screening test can often lead to substantial overdiagnosis.^9-11^

Although prospective studies of multicancer early detection tests have been conducted,^3,4^ none have been able to assess the potential for overdiagnosis due to a lack of comparator. While the upcoming NHS-Galleri trial results may provide estimates of overdiagnosis,^5^ several years of follow-up are required to provide reliable estimates of overdiagnosis. In cases such as these where empirical evidence is lacking, modeling studies are the only way to provide early estimates of the potential population-level impacts of screening. Prior modeling studies of multicancer screening tests have mostly focused on the potential mortality benefits of multicancer screening or harms from false positives, but not the potential for overdiagnosis.^12-15^

The objective of the current study was to assess the potential number of overdiagnoses and false positives that a multicancer early detection screening program in older adults could generate if it were implemented at the population level, using the setting of Canada as a case study.

## Materials & Methods

We expanded a previously developed microsimulation model (McGill Multicancer Model), which is a model of the health trajectories and survival of cancer cases. Previous model versions (1.0-2.0) were developed to assess the impact of delays in cancer diagnosis and treatment caused by the COVID-19 pandemic.^16,17^ For version (3.0) we expanded the model to include multicancer screening. A detailed description of the model is available as an online appendix.^18^ The sections below summarize the model structure and screening functionalities relevant to the current analysis.

### Cancer natural history & epidemiology

The model reproduces the cancer incidence and survival outcomes at 25 different sites by sex, age (0-105 years), and TNM stage based on Canadian cancer incidence and survival statistics.^19-25^ New cancer diagnoses are generated during each model 2-week time step based on sex- and age-specific incidence rates; historical incidence rates are used for 2000-2019, and forecasted incidence rates with exponential smoothing are used post-2019.^26^ An initial diagnosis date is established for each cancer during the time step. This is the clinical diagnosis date in absence of screening with a multicancer detection test, with diagnosis assumed to be due to either symptomatic detection, incidental detection, or detection through other single-site screening programs. For each cancer, a preclinical screen-detectable period is sampled prior to the clinical diagnosis date. The screen-detectable period is the period when the cancer is latent and asymptomatic but would be potentially detectable through screening. Due to the uncertainty regarding the duration of the screen-detectable period for most cancers, we assumed in the base case that all cancer sites have the same screen-detectable period. This duration is assumed to follow a truncated Weibull; the distribution parameters were varied in sensitivity analyses to assess the impact of different screen-detectable periods. Two death dates are selected for each cancer: (1) a cancer death date, based on net survival by cancer site, age, sex, and stage at diagnosis,^20-24^ and (2) an other cause death date, based on life tables by age and sex.^27^ The earliest of the two death dates determines whether a person dies from their cancer (Figure 1A) or dies from other causes first (Figure 1B).

**Figure 1.**
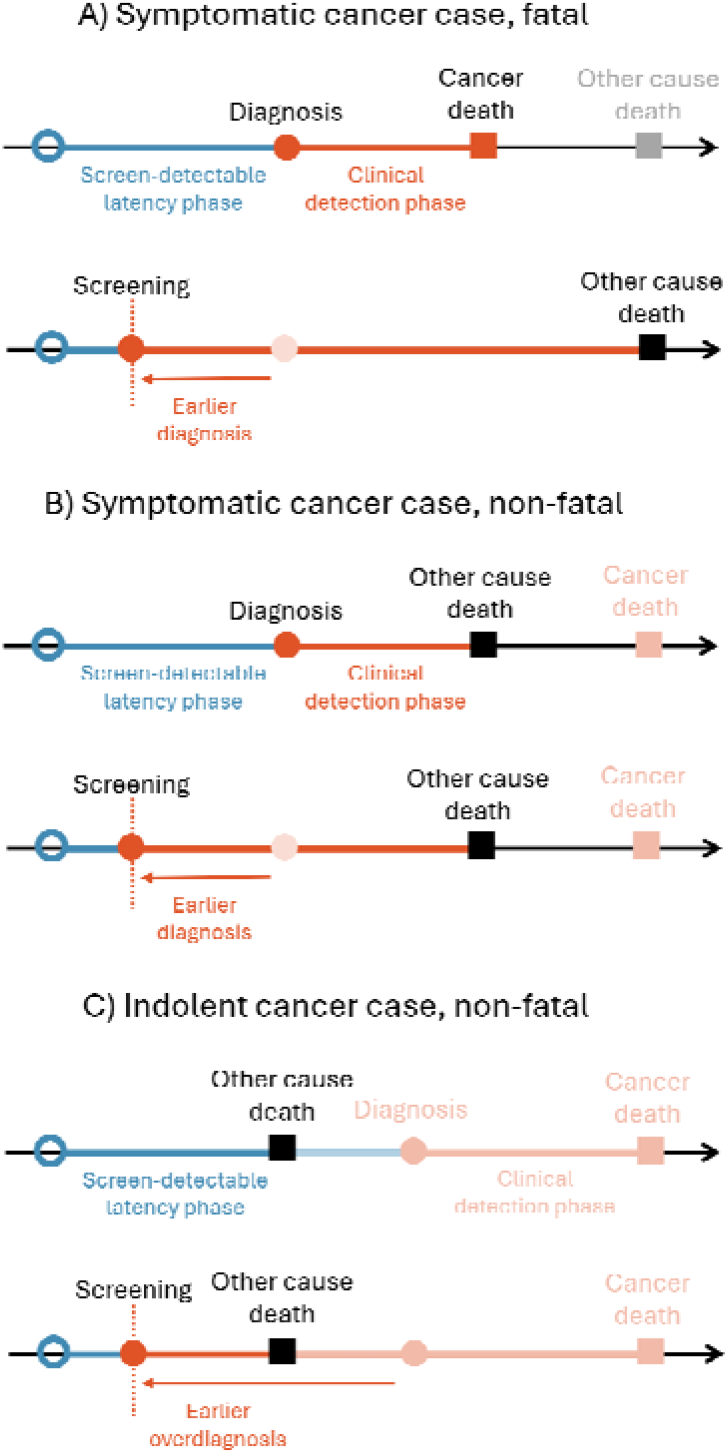
Conceptual diagram of cancer natural history in model. (A) Symptomatic cancers are those which would normally be detected due to symptoms or through incidental findings; for cases that would have died from their cancer, earlier detection through screening can potentially delay their cancer death, allowing them to die later from other causes. (B) Symptomatic cancers might also die from causes other than their cancer due to successful treatment or competing mortality risks; it is assumed earlier detection of these cancers would not affect their death date from other causes, so screening does not provide a mortality benefit for these cancers. These cancers are not considered to be overdiagnoses as they would have been diagnosed in absence of screening. (C) Indolent cancers are those which would not normally be diagnosed within a person’s lifetime; they have a death date from other causes sampled prior to their cancer diagnosis from symptomatic or incidental detection. These cancers can be detected through screening and are then counted as screening overdiagnoses, as they would not have been diagnosed without screening.

### Multicancer screening program

We simulated a multicancer screening program with a single test targeting multiple cancer sites starting in the year 2025. Since some tests are purported to detect cancers at over 50 different sites,^1^ we assumed that all 25 cancer sites in the model could potentially be detected by the test. The multicancer screening program is assumed to be additional to pre-existing single-cancer screening programs (breast, colorectal, cervical, lung). Screen-detected cancers, test results, and overdiagnoses reported in the results refer exclusively those detected through the multicancer screening program.

We assumed a multicancer screening program between ages 50-75 every 2-5 years. Screening intervals were based on cancer screening programs in Canada, which generally recommend 2-year (breast) to 5-year (cervical) screening intervals. A screening coverage parameter determines the probability of undergoing screening during screen-eligible ages. If a person is screened during a cancer’s screen-detectable period, then the probability of screen-detection of the cancer is based on test sensitivity. Given multicancer early detection tests have poorer sensitivity to earlier stage cancers,^28,29^ we assumed sensitivity increases with cancer stage. We assumed that all screen-positive cancers would undergo a diagnostic workup that would identify the underlying cancer.

### Screening Overdiagnosis & False Positives

Screening overdiagnosis is defined as the diagnosis of a cancer through screening which would not otherwise have been diagnosed during that person’s lifetime.^7^ The model simulates a pool of indolent cancers that would normally remain undiagnosed in absence of screening; these are individuals whose sampled death date from other causes is prior to their sampled cancer clinical diagnosis date (Figure 1C). These indolent cancers are flagged by the model as overdiagnoses if they are diagnosed through screening. Indolent cancers are assumed to be progressive lesions that would have eventually been diagnosed had the person lived longer. The model does not include non-progressive indolent lesions.

The incidence rate of indolent cancers by site (*i*), age (*a*), and sex (*s*) is modeled by multiplying observed cancer incidence rates by a multiplier (*θ*_*i,a,s*_), termed the indolence rate fraction:

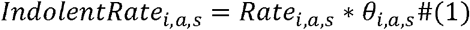

The indolence rate fraction is estimated based on the difference between cumulative risks of cancer in presence and in absence of competing mortality risks over an assumed latency period *x*. The numbers of expected diagnosed cancers at age *a* and at age *a*+*x* (*μ*_*i,a,s*_, *μ*_*i, a*+*x,s*_) are calculated by multiplying the cancer incidence rate (*Rate*_*i,a,s*_) by the person-years at risk (*PY*_*a,s*_) from life tables:

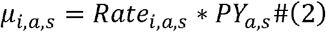

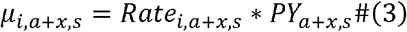

The number of latent cancer cases present at age *a* that would be diagnosed at age *a*+*x* in absence of competing mortality risks (*l*_*i,a,s*_) is:

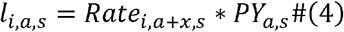

The difference between the expected number of cases in presence and in absence of competing risks is the number of indolent cases that will never be diagnosed due to a person dying before their cancer diagnosis (*o*_*i,a,s*_), which is used to calculate the indolence rate fraction (*θ*_*i,a,s*_):

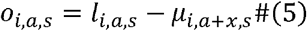

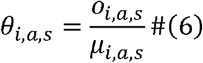

The indolence rate fraction strongly increases with age as latent cancers become increasingly less likely to be clinically diagnosed due to high competing mortality risks (Figure S1).

False positives are calculated based on the population size of age *a* and sex *s* (*Population*_*a,s*_), the screening coverage (*ϑ*_*a,s*_), the number of screen-detectable cancer cases in the population *n*_*a,s*_, and the screening test specificity (*Sp*):

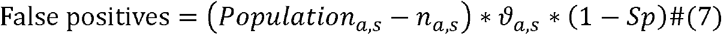

### Analyses

#### Cross-sectional analyses

The main outcomes of interest are the yearly number of screen-detected cancers, overdiagnosed cancer cases detected through screening, and test positive predictive value (PPV), the proportion of test positives that are true positives. We modeled scenarios where we varied the duration of the screen-detectable period, test sensitivity, test specificity, and screening interval to assess how each of these parameters influences potential for overdiagnosis. We assumed in analyses the same sensitivity, specificity, and latency periods across all cancer sites. The objective was not to model each cancer site in detail, but rather to cover a broad range of plausible scenarios from which to draw general conclusions about potential for overdiagnosis. The duration of the screen-detectable period is the most uncertain parameter. We examined scenarios with a 2- and 5-year maximum screen-detectable periods (most individual lead times being shorter due to sampling). The maximum screen-detectable period is used as the *x* parameter for calculating the indolent rate fractions. We also varied recommended screening intervals between 2 and 5 years, with an assumed 80% screening coverage. We assumed test sensitivities and specificities based on a previous study suggesting reasonable expectations for a multicancer test performance could be ∼40% sensitivity for stage I cancers and ∼80% sensitivity for stage III+ cancers.^28^ We also examined scenarios with lower test sensitivity and specificity. Model scenarios are summarized in Table 1. Each scenario was run 10 times to assess stochastic variation in predictions. The median (minimum-maximum range) of predictions is presented in results.

**Table 1.** Assumed model scenarios for multicancer test performance and screening program in base case scenario.

| Parameter | Across all scenarios |  |
| --- | --- | --- |
| Screening Coverage | 80% |  |
| Screening ages | 50-75 years |  |
| Varied by scenario | Base case scenario | Alternative scenarios |
| Test sensitivity by stage (all cancer sites)<sup>a</sup> |  |  |
| Stage I | 40% | 20% |
| Stage II | 50% | 30% |
| Stage III | 60% | 40% |
| Stage IV | 80% | 60% |
| Unknown | 50% | 30% |
| Test Specificity | 99.1% | 97% |
| Max screen-detectable period<sup>b</sup> | 5 years | 2 years |
| Screen-detectable period Weibull shape parameter | 5.5 | 2.5 |
| Screen-detectable period Weibull scale parameter | 4.3 | 1.6 |
| Median (Interquartile range) screen-detectable period | 4.0 (3.4-4.6) | 1.4 (1.0-1.8) |
| Screening interval | 5 years | 2 years |

<sup>a</sup>Sensitivity for unstageable cancer sites (e.g. leukemia) is assumed to be 50/30% in the base case/alternative scenarios.
<sup>b</sup>The maximum screen-detectable period is the upper limit of the truncated Weibull distribution, and is used as the latency period for generating indolent cancers.

#### Cohort analyses

Cumulative risks of outcomes over a lifetime of screening were generated by simulating a population of 100,000 persons starting at age 45 until their deaths. Because the model does not track multiple primary tumors within the same person, a correction factor was applied to calculate cumulative risk accounting for the probability of a previous cancer diagnosis. Detailed formulas for this correction factor, as well as model validation, can be found in the technical appendix.^18^

## Results

The introduction of a multicancer screening program was predicted to lead to a short-term increase in diagnosed cancer incidence among the 50-79 years age groups targeted by screening (Figure 2). However, in the scenario with a 5-year screening interval and 5-year screen-detectable period, incidence rates were predicted to eventually return to pre-screening levels over the long-term. In scenarios with a 2-year screening interval and screen-detectable period, there was a shift in the distribution of age at diagnosis of cancer incidence, with incidence rates increasing in age groups with 3 screening opportunities (50-54, 60-64, 70-74 years) and rates decreasing in age groups with only 2 screening opportunities (55-59, 65-69). There were also lower incidence rates in the age group of 75-79 years due to the last screening occurring at age 74 in scenarios with 2-year screening intervals. These results suggest the introduction of a multicancer screening program is unlikely to lead to a permanent increase in cancer incidence due to overdiagnosis, but that the age distribution of cancer cases might be impacted by the screening interval if screening opportunities are unequally distributed across age groups.

**Figure 2.**
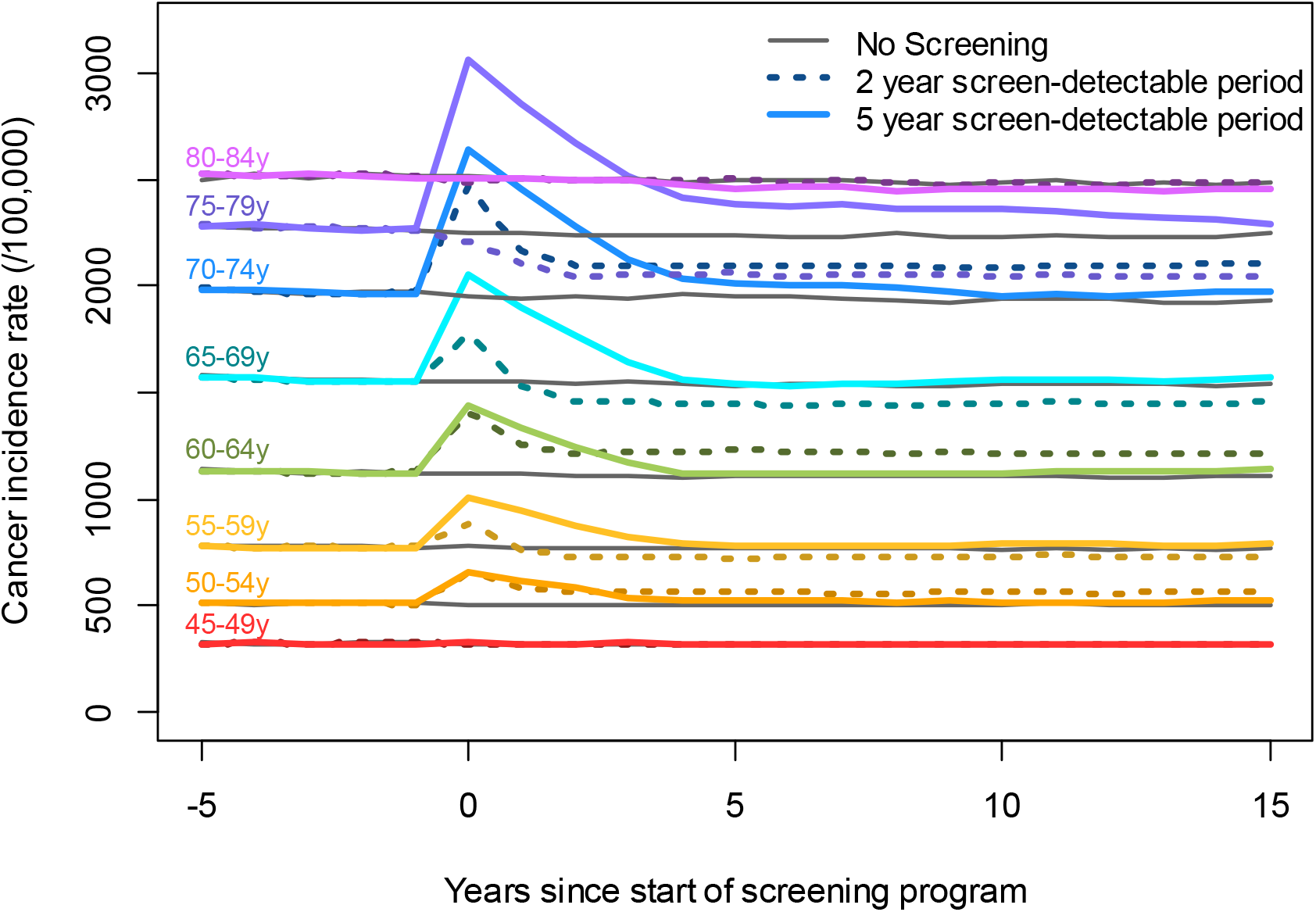
Overall cancer incidence rate (/100,000 person-years) by age group in a no screening scenario (grey lines) and in screening scenarios (colored lines), by years since program implementation. The solid-colored lines show predictions for a scenario with an assumed 5-year screen-detectable period and a 5-year screening interval. The dashed-colored lines show predictions for a scenario with an assumed 2-year screen-detectable period and a 2-year screening interval. Screening scenarios assume 40/50/50/80% sensitivity for stage I/II/III/IV and 99.1% specificity. Ages 50-75 are targeted for screening; age groups 45-49, 80-84 years are included for comparison. Results are the median of 10 model runs.

In the base case scenario of a multicancer screening program for 50–75-year-olds tested every 5 years (Table 2), by 2035 there would be approximately 302,233 (range across stochastic runs 301,606-303,907) cancers diagnosed each year across all ages in Canada, of which 70,277 (69,760-70,327) would be screen-detected by the multicancer screening test. Of the screen-detected cancers, 4,192 (4,033-4,229) or 6.0% (5.7-6.0%) were predicted to be screening overdiagnoses, cancers that would not have been clinically detected within the person’s lifetime had the person not screened. The probability of overdiagnosis strongly increased with screening age, with 1.1% (0.9-1.2%) of screen-detected cancers predicted to be overdiagnoses at ages 50-54 years, and 10.8% (10.5-11.1%) of screen-detected cancers predicted to be overdiagnoses at ages 75-79 years. The predicted proportion of overdiagnosis in screen-detected cancers was higher in males (7.3%) than in females (4.5%), and varied by cancer site, ranging from 2.4% (0.0-4.8%) for testicular cancer to 7.7% (5.5-8.0%) for esophageal cancer.

**Table 2.** Yearly screen-detected cancers and overdiagnoses at each age by cancer site in base case scenario 10 years after the start of the screening program.

|  | Screen-detected | Overdiagnoses | % Overdiagnosis |
| --- | --- | --- | --- |
|  | Median (Min-max runs) | Median (Min-max runs) | Median (Min-max runs) |
| All | 70,277 (69,760-70,327) | 4,192 (4,033-4,229) | 6.0 (5.7-6.0) |
| By Sex |  |  |  |
| Female | 33,036 (32,767-33,069) | 1,473 (1,397-1,529) | 4.5 (4.2-4.6) |
| Male | 37,248 (36,944-37,380) | 2,718 (2,602-2,791) | 7.3 (7.0-7.5) |
| By Age Group |  |  |  |
| 50-54 years | 4,904 (4,802-5,082) | 52 (42-60) | 1.1 (0.9-1.2) |
| 55-59 years | 6,988 (6,807-7,061) | 122 (95-137) | 1.7 (1.4-2.0) |
| 60-64 years | 9,301 (9,045-9,368) | 247 (242-292) | 2.7 (2.6-3.1) |
| 65-69 years | 12,783 (12,454-12,952) | 540 (508-579) | 4.2 (4.0-4.5) |
| 70-74 years | 17,029 (16,869-17,150) | 1,096 (1,062-1,175) | 6.4 (6.2-6.9) |
| 75-79 years | 19,275 (18,993-19,441) | 2,084 (2,017-2,134) | 10.8 (10.5-11.1) |
| By Cancer Site<sup>a</sup> |  |  |  |
| Thyroid | 1,186 (1,130-1,229) | 44 (38-55) | 3.7 (3.2-4.6) |
| Testis | 42 (30-52) | 1 (0-2) | 2.4 (0.0-4.8) |
| Prostate | 9,074 (8,919-9,111) | 636 (601-680) | 7.0 (6.6-7.5) |
| Melanoma | 2,607 (2,570-2,657) | 148 (139-166) | 5.7 (5.3-6.4) |
| Breast | 7,439 (7,321-7,548) | 304 (279-325) | 4.1 (3.8-4.4) |
| Hodgkin Lymphoma | 177 (157-224) | 10 (6-12) | 5.6 (3.4-6.8) |
| Uterus | 2,178 (2,140-2,292) | 95 (69-99) | 4.4 (3.2-4.5) |
| Bladder | 3,091 (2,981-3,193) | 221 (213-263) | 7.1 (6.9-8.5) |
| Cervix | 237 (201-265) | 8 (4-12) | 3.4 (1.7-5.1) |
| Kidney & Renal Pelvis | 2,246 (2,222-2,308) | 125 (111-153) | 5.6 (4.9-6.8) |
| Non-Hodgkin Lymphoma | 3,654 (3,602-3,750) | 231 (206-245) | 6.3 (5.6-6.7) |
| Colorectal | 5,736 (5,676-5,795) | 336 (296-363) | 5.9 (5.2-6.3) |
| Oral | 4,346 (4,234-4,440) | 244 (223-264) | 5.6 (5.1-6.1) |
| Larynx | 470 (455-504) | 32 (29-39) | 6.8 (6.2-8.3) |
| Leukemia | 1,733 (1,672-1,834) | 116 (102-135) | 6.7 (5.9-7.8) |
| Central Nervous System | 27 (19-43) | 1 (0-1) | 3.7 (0.0-3.7) |
| Multiple Myeloma | 1,104 (1,002-1,123) | 71 (61-78) | 6.4 (5.5-7.1) |
| Ovary | 900 (866-963) | 36 (34-52) | 4.0 (3.8-5.8) |
| Stomach | 1,496 (1,432-1,550) | 108 (91-116) | 7.2 (6.1-7.8) |
| Lung | 10,779 (10,688-10,807) | 710 (636-719) | 6.6 (5.9-6.7) |
| Brain | 720 (662-755) | 37 (25-44) | 5.1 (3.5-6.1) |
| Liver | 1,304 (1,267-1,337) | 86 (63-105) | 6.6 (4.8-8.1) |
| Esophagus | 932 (896-991) | 72 (51-75) | 7.7 (5.5-8.0) |
| Pancreas | 2,475 (2,455-2,545) | 159 (156-191) | 6.4 (6.3-7.7) |
| Other | 6,190 (6,103-6,271) | 340 (319-373) | 5.5 (5.2-6.0) |

<sup>a</sup> Cancer sites are ordered by decreasing 5-year relative survival probability in Canada, with the exception of Other cancers which is presented last

Results of scenario analyses varying the screen-detectable latency period, screening interval, test sensitivity, and test specificity are presented in Table 3. The screen-detectable latency period was the parameter that most strongly impacted the number and proportion of cancer overdiagnoses. Only a few hundred overdiagnosed cancers were predicted in scenarios where cancers have a 2-year maximum screen-detectable period, compared with thousands predicted in scenarios where cancers have a 5-year maximum screen-detectable period. The proportion of overdiagnosis in screen-detected cancers ranged from 2.1% (2.0-2.3%) in scenarios with a 2-year maximum latency period, to 6.0% (5.7-6.0%) in scenarios with a 5-year maximum latency period. The test sensitivity and screening intervals did not much impact the proportion of screen-detected cancers that were overdiagnoses. Test specificity did not much impact either the number or proportion of predicted overdiagnoses, but had a strong impact on the number of false positives and consequently the PPV of the screening test, which ranged from lower values of 15.9% (15.8-16.0%) in scenarios assuming lower (97%) test specificity and shorter screen-detectable periods, up to 77.6% (77.4-77.6%) in scenarios assuming higher (99.1%) test specificity and longer screen-detectable periods.

**Table 3.** Yearly screening overdiagnoses between ages 50-75y under different test performance & program parameter assumptions, 10 years after the start of the screening program.

| Max screen-detectable<br>period | Screening<br>interval | Sensitivity by<br>stage I/II/III/IV | Specificity | True<br>Positives/<br>Screen-detected<br>cancers (n) | False<br>positive<br>(n) | PPV<sup>a</sup> | NNS<sup>a,b</sup> | Overdiagnoses |  |
| --- | --- | --- | --- | --- | --- | --- | --- | --- | --- |
|  |  |  |  |  |  |  |  | n<sup>a</sup> | Proportion of<br>screen-detected<br>cancers (%)<sup>a</sup> |
| Shorter (2y) | Shorter<br>(2y) | Lower<br>(20/30/40/60%) | Lower (97%) | 29,042 | 153,279 | 15.9 (15.8-16) | 179 (180-178) | 615 (570-658) | 2.1 (2.0-2.3) |
|  |  |  | Higher (99.1%) | 28,984 | 45,984 | 38.7 (38.5-38.9) | 179 (180-177) | 631 (577-654) | 2.2 (2.0-2.3) |
|  |  | Higher<br>(40/50/60/80%) | Lower (97%) | 46,144 | 153,276 | 23.1 (22.9-23.2) | 113 (114-112) | 984 (934-1,056) | 2.1 (2.0-2.3) |
|  |  |  | Higher (99.1%) | 45,835 | 45,984 | 49.9 (49.8-50) | 113 (114-113) | 981 (903-1,036) | 2.1 (2.0-2.3) |
|  | Longer (5y) | Lower<br>(20/30/40/60%) | Lower (97%) | 14,848 | 70,199 | 17.5 (17.3-17.6) | 160 (162-159) | 366 (339-382) | 2.5 (2.3-2.6) |
|  |  |  | Higher (99.1%) | 14,836 | 21,060 | 41.3 (41.2-41.7) | 161 (162-158) | 360 (353-397) | 2.4 (2.4-2.7) |
|  |  | Higher<br>(40/50/60/80%) | Lower (97%) | 23,552 | 70,192 | 25.1 (24.9-25.4) | 101 (102-100) | 567 (537-584) | 2.4 (2.3-2.5) |
|  |  |  | Higher (99.1%) | 23,627 | 21,059 | 52.9 (52.7-53.1) | 101 (102-100) | 575 (551-606) | 2.4 (2.3-2.6) |
| Longer (5y) | Shorter<br>(2y) | Lower<br>(20/30/40/60%) | Lower (97%) | 78,382 | 148,957 | 34.5 (34.2-34.6) | 66 (67-66) | 4,066 (3,947-4,144) | 5.2 (5.0-5.3) |
|  |  |  | Higher (99.1%) | 78,183 | 73,810 | 53.4 (43.1-63.7) | 108 (108-108) | 4,059 (3,945-4,159) | 5.2 (5.0-5.3) |
|  |  | Higher<br>(40/50/60/80%) | Lower (97%) | 114,231 | 149,490 | 43.3 (43.3-43.4) | 45 (46-45) | 5,856 (5,759-5,973) | 5.1 (5.0-5.2) |
|  |  |  | Higher (99.1%) | 114,234 | 44,848 | 71.8 (71.8-71.9) | 45 (46-45) | 5,938 (5,760-6,090) | 5.2 (5.0-5.3) |
|  | Longer (5y) | Lower<br>(20/30/40/60%) | Lower (97%) | 44,362 | 67,748 | 39.6 (39.4-39.8) | 54 (54-53) | 2,608 (2,538-2,657) | 5.9 (5.7-6.0) |
|  |  |  | Higher (99.1%) | 44,400 | 20,324 | 68.6 (68.5-68.7) | 54 (54-53) | 2,616 (2,541-2,698) | 5.9 (5.7-6.1) |
|  |  | Higher<br>(40/50/60/80%) | Lower (97%) | 70,318 | 67,708 | 50.9 (50.8-51.1) | 34 (34-34) | 4,120 (4,061-4,247) | 5.9 (5.8-6.0) |
|  |  |  | Higher (99.1%) | 70,277 | 20,313 | 77.6 (77.4-77.6) | 34 (34-34) | 4,192 (4,033-4,229) | 6.0 (5.7-6.0) |

NNS=Number needed to Screen; PPV=Positive Predictive Value of screening test
<sup>a</sup> Parentheses indicate the minimum-maximum over 10 model runs
<sup>b</sup> Number needed to screen to detect one cancer

In cohort analyses, the cumulative probability of being diagnosed with cancer between ages 45-80 years was 32.1% (32.0-32.3%); assuming a person undergoes a multicancer screening every 5 years between ages 50-75, the cumulative probability by age 80 of a positive screening test result was 18.2% (18.1-18.3%); of having a screen-detected cancer was 14.4% (14.3-14.5%); and of being overdiagnosed with a cancer through screening was 0.9% (0.9-1.0%). These cumulative probabilities were slightly higher in males than females (Figure 3).

**Figure 3.**
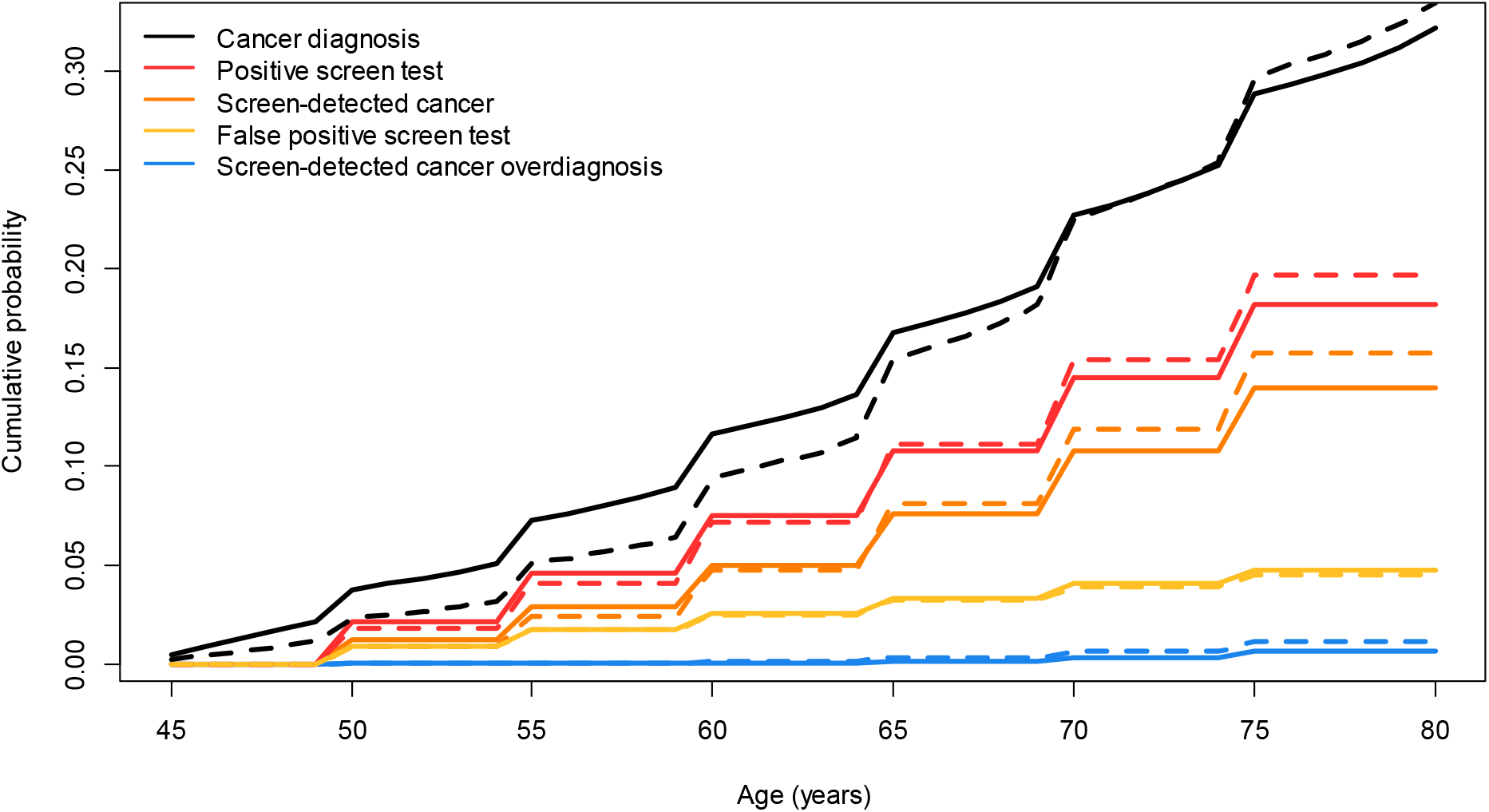
Cumulative probability between ages 45-80 of a cancer diagnosis (black), receiving a positive screen test (red), having a screen-detected cancer (orange), receiving a false positive test result (yellow) and of being overdiagnosed with a cancer through screening (blue) by sex. Solid lines are for females and dashed lines are for males. The cohort is assumed to screen every 5 years between ages 50-75 years (100% adherence), cancers have an assumed 5 years maximum screen-detectable period, test specificity 99.1%, test sensitivity 40/50/60/80% for stages I/II/III/IV. Results are the median of 10 model runs.

## Discussion

In this study, we modeled the potential impact of a multicancer screening program between ages 50-75 years and found a relatively low rate of overdiagnosis proportional to the total number of screen-detected cancers. Based on plausible ranges of test performances, a reasonable expectation would be that some 2.1-6.0% of all screen-detected cancers with a multicancer screening test could be overdiagnoses. The proportion of overdiagnosis strongly increased with age, going from only 1% at age 50 to over 10% of all screen-detected cancers by age 75. Differences across cancer sites were influenced by average age at onset, as overdiagnosis fractions tended to be lower for cancers with earlier onset ages such as testicular cancer, and higher for cancers with later onset ages such as bladder cancer. Apart from age, the most important factor determining the potential for overdiagnosis was not test sensitivity or specificity, but rather the screen-detectable period of the cancers targeted by the screening test. The longer the average screen-detectable period across cancers, the higher the proportion of screen-detected cancers that will be indolent cancers that would not have been diagnosed within that person’s lifetime. As expected, the most important factor influencing false positive results relative to true positive (PPV) was the test specificity.

The screen-detectable period of most cancers is highly uncertain. Due to this uncertainty, we opted to assess scenarios with shorter and longer screen-detectable periods rather than make unverifiable assumptions for specific screen-detectable periods by cancer site. The 2.1-6.0% range is meant to represent a plausibility interval for the overdiagnosis fraction given a reasonable range of screen-detectable periods. We expect the upcoming results of the NHS-Galleri trial to be enlightening regarding the potential distribution of screening lead times,^5^ which will likely vary by cancer site. These data would allow us to implement more informed site-specific screen-detectable periods in future analyses. The screen-detectable period may be longer for cancers sites with high cell turnover such as colorectal cancer, and shorter for cancers with low cell turnover such as breast cancer; for example, a previous model of tumour growth and ctDNA shedding rates has estimated a probable lead time on the order of 300-450 days for lung cancer.^30^ A cohort study found that methylated DNA cancer signals may in some cases be detectable up to 3 years prior to diagnosis.^31^ As a comparison point, the estimated mean screen-detectable periods for lung, breast, colorectal cancer screening tests have generally ranged from 1-6 years.^32-36^ Our scenarios are therefore consistent with what is plausible for cell-free DNA biomarkers and with screen-detectable periods estimated for prior cancer screening tests.

Our estimates of overdiagnosis can be contextualized by comparison with lung cancer screening. Overdiagnosis fractions estimated from low-dose computed tomography screening trials have ranged from 0% to 67% of all screen-detected lung cancers.^37-39^ Some of the variation may be attributable to differences in follow-up across trials; follow-up needs to extend long enough to cover all screening lead times to provide valid estimates of overdiagnosis. For example, while initial estimates from the NLST trial were that 18.5% of screen-detected cancers were overdiagnoses after 6.3 years,^36^ extended follow-up led to a reduced estimate of only 3% excess cancers after 11.3 years.^40^ This is likely due to a subset of lung cancers with very long lead times ,^37^ with many of the cancers initially thought to be overdiagnoses being in fact screen-detected cancers with very long lead times of over 10 years. As the lead times with ctDNA are believed to be much shorter,^30^ it is consequently unsurprising that we estimated lower overdiagnosis fractions (e.g. 6.6% for lung cancer) when assuming maximum screen-detectable periods of 2-5 years.

What constitutes an acceptable false positive risk in the context of a multicancer screening program may depend on health system capacity. Our model predicts that a population-based multicancer screening program could be expected to have relatively high PPVs (15.9-77.6%), in line with results reported in clinical studies of these tests in highly selected clinical populations^3,4^ and other modeling projections.^13^ These high PPVs can be explained by the high prevalence of underlying cancer in 50-75 year olds. While these PPVs are higher than those reported for other single-site cancer screening programs,^41,42^ the potential harms and costs from a false positive multicancer screening test result are also higher. Positive multicancer test results risk launching patients on a “diagnostic odyssey” to identify the putative cancer site and rule out the presence of any cancer in absence of confirmatory findings. While studies have shown it is possible to implement diagnostic pathways in test positives (e.g. full-body imaging),^3,4^ it remains unclear to what extent these approaches are scalable and cost-effective at the population-level. For these reasons, it may be justifiable to require a higher PPV for implementation of multicancer screening due to the more complex diagnostic workup required.

Our analysis presents two important limitations. The first is that we only considered overdiagnosis from progressive lesions. There is however ample evidence that non-progressive indolent lesions exist, and that the detection of these lesions has contributed to substantial overdiagnosis of breast and prostate cancers through screening.^7,9-11^ It has been hypothesized that the detection of non-progressive lesions would be much lower with multicancer early detection tests, because ctDNA is a prognostic marker linked to cancer mortality and tumour burden.^8^ Multicancer early detection tests are therefore hypothesized to mostly detect progressive cancers that would eventually lead to a lethal tumour burden. Even so, our estimates of potential overdiagnosis are likely conservative. Secondly, the model does not track multiple primary tumours per person. It is unclear how the presence of synchronous tumours at different sites would affect screening test results and subsequent overdiagnosis. However, as only <2% of cancers present as multiple synchronous primaries,^43^ it is unlikely this has substantially impacted results.

## Conclusion

Previous cancer screening programs have demonstrated that substantial overdiagnosis can occur with cancer screening, and overdiagnosis has played a major role in shaping recommendations against screening with certain tests. Overdiagnosis is consequently one of the most important harms to consider as we start to think about potentially screening with multicancer early detection test. Our study suggests that the proportion of screen-detected cancers that would be overdiagnoses would likely not be very high, and that a multicancer screening program would likely not lead to a substantial long-term increase in cancer incidence from overdiagnosis as has been seen with previous cancer screening tests.^9,11^ While a multicancer screening program could potentially lead to the earlier detection of many cancers, as earlier detection does not necessarily lead to decreased cancer mortality, we plan in future analyses to examine potential impacts on mortality outcomes. As the risk of overdiagnosis strongly increases with age, there would likely be an upper age limit above which the harms of multicancer screening would outweigh the mortality benefits. This upper age limit will likely vary by population depending on cancer incidence, life expectancy, and tolerance for risk.

## Acknowledgements

Computations were made on the Rorqual supercomputer, managed by Calcul Québec and the Digital Research Alliance of Canada. The operation of this supercomputer is funded by the Digital Research Alliance of Canada, le ministère de l’Économie, de l’Innovation et de l’Énergie du Québec (MEIE) and le Fonds de recherche du Québec (FRQ). We thank Ruth Etzioni for her comments on early drafts of this paper.

## Data Availability

Model predictions and codes to generate results are available on the Borealis repository at https://doi.org/10.5683/SP3/KMWOYZ. Model technical documentation is available at https://doi.org/10.5683/SP2/REMSZ6.

## Ethics Statement

The current study did not involve human participants and is based on publicly available information. This study is therefore exempt from ethical approval.

