## Supplemental Figure 1 for "Assessing potential harms from screening overdiagnosis and false positives with multicancer early detection tests"

Cancer Incidence (/year)

Indolence rate fraction

| Age | Incidence of indolent cancers | Incidence of clinically detected cancers | Indolence rate fraction |
| --- | --- | --- | --- |
| 0 to 4 years | 0 | 460 | 0 |
| 5 to 9 years | 2 | 281 | 0.00711744 |
| 10 to 14 years | 0 | 278 | 0 |
| 15 to 19 years | 1 | 519 | 0.00192678 |
| 20 to 24 years | 5 | 924 | 0.00541126 |
| 25 to 29 years | 2 | 1500 | 0.00133333 |
| 30 to 34 years | 8 | 2498 | 0.00320256 |
| 35 to 39 years | 14 | 3866 | 0.00362131 |
| 40 to 44 years | 35 | 5253 | 0.00666286 |
| 45 to 49 years | 50 | 7772 | 0.00643335 |
| 50 to 54 years | 126 | 12437 | 0.0101311 |
| 55 to 59 years | 320 | 21450 | 0.0149184 |
| 60 to 64 years | 788 | 29154 | 0.0270289 |
| 65 to 69 years | 1618 | 34182 | 0.0473349 |
| 70 to 74 years | 2705 | 35836 | 0.0754828 |
| 75 to 79 years | 4273 | 27714 | 0.154182 |
| 80 to 84 years | 5448 | 20362 | 0.267557 |
| 85 to 89 years | 6185 | 13487 | 0.45859 |
| 90 to 94 years | 5891 | 6101 | 0.965579 |
| 95 to 99 years | 3904 | 1666 | 2.34334 |
| 100 to 104 years | 1334 | 247 | 5.40081 |

Figure S1. Cancer incidence and indolence rate fraction by age in 2020 for all cancers combined, both sexes combined when assuming a 5-year latency/screen-detectable period. The indolence rate fraction can be interpreted as the ratio of indolent screen-detectable cancers arising at each age relative to the observed clinical cancer incidence at that age. For example, in 2020 there are 0.27 indolent undiagnosed but screen-detectable cancers arising for every clinically diagnosed cancer in 80–84-year-olds. These indolent cancers would not normally be diagnosed as the person is expected to die within the next 5 years prior to the clinical cancer diagnosis, unless screening detects the cancer early.
